# Educational attainment in hemorrhagic stroke severity and functional outcome

**DOI:** 10.1101/2023.02.21.23286280

**Authors:** Nirupama Yechoor, Pamela Rist, Alena Ganbold, Christina Kourkoulis, Samantha Mora, Ernst Mayerhofer, Livia Parodi, Christopher D. Anderson, Jonathan Rosand

**Affiliations:** Division of Neurocritical Care, Massachusetts General Hospital, Boston, MA, USA; Henry and Allison McCance Center for Brain Health, Massachusetts General Hospital, Boston, MA, USA; Division of Preventive Medicine, Department of Medicine, Brigham and Women’s Hospital and Harvard Medical School, Boston, MA, USA; Department of Epidemiology, Harvard T.H. Chan School of Public Health, Boston, MA, USA; Department of Neurology, Massachusetts General Hospital, Boston, MA, USA; Center for Genomic Medicine, Massachusetts General Hospital, Boston, MA, USA; Program in Medical and Population Genetics, Broad Institute of Harvard and the Massachusetts Institute of Technology, Cambridge, MA, USA; Department of Neurology, Brigham and Women’s Hospital, Boston, MA, USA

**Keywords:** Intracerebral hemorrhage, social determinants of health, education, modifiable risk factors, stroke severity, stroke outcomes

## Abstract

**Introduction:** While stroke education is a hallmark of clinical care in primary stroke centers, the educational attainment of patients is rarely considered. The objective of this study is to determine whether educational attainment, in addition to traditional risk factors, is associated with intracerebral hemorrhage (ICH) severity and functional outcomes.

**Methods:** Subjects were enrolled in a prospectively ascertained cohort of patients with primary ICH from 1994 until 2020 at the Massachusetts General Hospital Neurosciences Intensive Care Unit. Data on social determinants of health (SDOH), past medical history for ICH risk factors, and ICH severity were obtained on admission. The primary outcome was stroke severity, calculated using the ICH score. The secondary outcome was 12-month modified Rankin Score (mRS).

**Results:** Of 2,539 patients eligible for analyses, 1,744 (69%) presented with mild ICH (ICH Score < 3) and 795 (31%) presented with severe ICH (ICH score ≥3). In a multivariable logistic regression analysis controlling for age, income, and a pre-stroke diagnosis of coronary artery disease, only educational attainment was associated with stroke severity, with patients with high school-only education more likely to present with a severe ICH compared to college diplomates (odds ratio 2.37, 95% CI 1.77, 3.19).

Additionally, those with high school-only education were less likely to recover to a good functional outcome (mRS<4) after ICH when compared to college diplomates (odds ratio 0.43, 95% CI 0.19, 0.93) when controlling for stroke severity, and pre-stroke diagnoses of hypertension and dementia.

**Discussion:** Pre-stroke educational attainment is an independent predictor of ICH severity and subsequent functional outcomes at 12-month post-stroke. Given the association with recovery, tailoring inpatient stroke education to level of educational attainment may serve to reduce the disparity in outcomes after hemorrhagic stroke. Educational attainment may act as both a social and a biological determinant, and could represent a modifiable risk factor for ICH survivors.

## Introduction

Healthy People 2030 highlights the important role of social determinants of health (SDOH) in impacting peoples’ health and wellbeing, and the crucial contribution of SDOH to health disparities^1,2^. This framework categorizes various SDOH into five major domains of Economic Stability, Education Access and Quality, Healthcare Access and Quality, Neighborhood Environment, and Social and Community Context. Notably, educational attainment has been previously studied as an independent risk factor for all-cause mortality and increased risk of cardiovascular and cerebrovascular disease^3^.

Given the importance of education in health outcomes, stroke education has naturally been a hallmark of clinical care in primary stroke centers. Further supporting the importance of education, previous research in ischemic stroke shows that low educational attainment increases the risk for incident stroke, stroke severity, and impact outcomes in stroke recovery^3-9^. Additionally, some studies have also found that educational attainment is a protective factor against stroke and is associated with improved cognitive recovery even after large ischemic strokes^6,7^. However, it is unknown if educational attainment impacts hemorrhagic stroke severity or recovery.

Despite increasingly standardized care for acute clinical management of intracerebral hemorrhage (ICH), disparate outcomes after ICH have been described^10-16^. Given prior literature suggesting a role for educational attainment impacting risk and recovery for stroke, we hypothesize that in addition to traditional clinical risk factors, such as hypertension, education will be associated with ICH severity and recovery. Further investigating this relationship could lead to novel interventions for secondary prevention that are more patient-focused to accommodate for differences in educational attainment. Additionally, understanding the relationship of pre-stroke education levels could lead to new strategies for primary prevention in those at highest risk for ICH. Therefore, this study aims to understand if educational attainment is associated with stroke severity on admission for ICH patients, in addition to investigating the association with long-term functional outcome.

## Methods

### Study Design

We used data from the prospectively collected Massachusetts General Hospital ICH cohort^10^. Inclusion criteria are age over 18 and admission to the MGH Neuroscience Intensive Care Unit from January 1994 until January 2020 with a clinical and radiographically confirmed diagnosis of primary ICH. Exclusion criteria are those patients with missing clinical data of stroke severity that could not have an ICH score calculated, and those with ICH secondary to traumatic injury, brain tumor, or a vascular malformation. Demographic data of age, sex, and zip code information were collected. To evaluate the impact of traditional risk factors on ICH severity, medical history collected included a diagnosis of hypertension (HTN) prior to admission, history of tobacco use, diabetes, coronary artery disease (CAD), atrial fibrillation, history of liver disease, diagnosis of dementia at time of admission, and history of prior hemorrhagic stroke. This study adhered to the STROBE observational cohort guidelines^17^.

### Social Determinants of Health

The Healthy People 2020 framework guided the selection of SDOH to include the 5 domains: 1) economic stability, 2) education, 3) social/community context, 4) neighborhood environment, and 5) access to healthcare. The following SDOH were collected for all patients: median income, highest level of education, marital status, race, and religion.

For median income, raw income was stratified into four groups: low-income representing <$40,000 U.S dollars (USD) annually, middle-income representing an annual salary of $40,000-$75,000 USD, high-income representing an annual salary of $75,000-$100,000, and a very high income representing an annual salary of >$100,000 USD annually.

Marital status was grouped by married, separated, single, and widowed. Race was self-stated and grouped by white, black, Asian/Pacific Islander, or multiple races and/or another race. Educational attainment was stratified as any high school education including high school diplomat, or college diplomat. Religions represented included Christian, Non-Christian, and Non-affiliated.

### Stroke Severity

Stroke severity served as the primary outcome. The ICH Score was chosen as the measure of stroke severity given it is widely used and captures GCS on admission, age, location and volume of hemorrhage, and presence of intraventricular hemorrhage (IVH). The ICH score is an ordinal categorical variable ranging from 0 to 6, which for this study was dichotomized into mild (0-2) and severe (≥ 3) stroke.

### Functional outcomes

Functional outcome was the secondary outcome, based on modified Rankin scale (mRS) score at 12 months. The measure of functional outcome was obtained by telephone follow-up by research staff to assess mRS, in addition to clinical notes available for review. For all analyses, mRS was dichotomized into good (mRS 0-3) and poor (4-6) outcome.

### Data Analysis

Analyses were performed in R and R Studio (www.r-project.org). First, we compared baseline characteristics, past medical history, and SDOH among those with mild stroke versus severe stroke using the Wilcoxon Rank Sum test and chi-square tests. Next, factors which were significantly associated with stroke severity in univariate analyses were then included in a logistic regression model. This approach was then repeated among those with functional outcome data at 12 months.

Less than 100 participants were missing information on income bracket, education, history of hypertension, history of diabetes, history of coronary artery disease, history of atrial fibrillation, history of dementia, history of prior hemorrhage and were excluded from analysis. More than 100 participants were missing for history of tobacco use and history of liver disease; these participants were assigned to a separate “missing” category and included in the analysis. Participants with missing information on race were included in the “other” category.

## Results

### Study population

Of the 3,186 patients in the MGH ICH cohort, 2,539 patients had complete stroke severity data available on admission. Of these patients, 1,744 (69%) of them presented with mild ICH (ICH Score less than 3) and 795 (31%) presented with severe ICH (ICH score 3 or greater). The median age of those with mild strokes was 73, compared with 78 for those with severe strokes (Table 1). Approximately 10% of the cohort had a prior hemorrhage and most participants had a diagnosis of hypertension (Table 2). Patients with a diagnosis of CAD were more likely to present with a severe stroke on admission (p-value = 0.003).

**Table 1:**
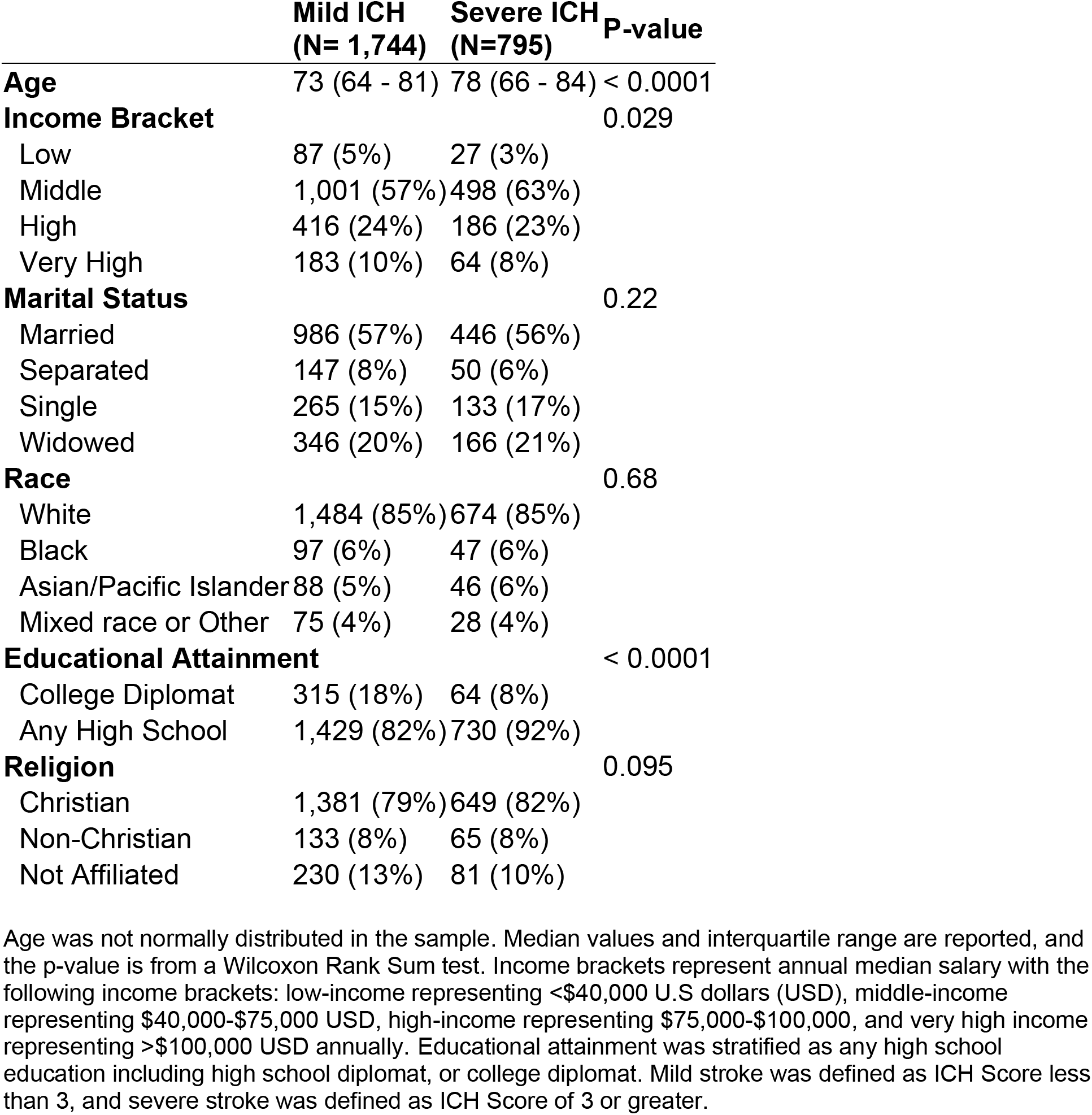
Social determinants of health and ICH severity

**Table 2:** Pre-ICH medical history stratified by severity

|  | **Mild ICH**<br>**(N=1,744)** | **Severe ICH**<br>**(N=795)** | **P-value** |
| --- | --- | --- | --- |
| **Hypertension** |  |  | 0.63 |
|  | 1,377 (79%) | 632 (79%) |  |
| **Sex** |  |  | 0.37 |
| Female | 800 (46%) | 380 (48%) |  |
| **Smoking** |  |  | 0.36 |
| Current | 216 (12%) | 74 (9%) |  |
| History | 606 (35%) | 222 (28%) |  |
| Never | 659 (38%) | 272 (34%) |  |
| Missing | 263 (15%) | 227 (29%) |  |
| **Diabetes** |  |  | 0.41 |
|  | 371 (21%) | 180 (23%) |  |
| **Coronary Artery Disease** |  |  | 0.003 |
|  | 349 (20%) | 201 (25%) |  |
| **Atrial Fibrillation** |  |  | 0.15 |
|  | 367 (21%) | 188 (24%) |  |
| **Liver Disease** |  |  | 1.0 |
| Yes | 59 (3%) | 26 (3%) |  |
| Missing | 202 (12%) | 109 (14%) |  |
| **Dementia** |  |  | 0.49 |
|  | 238 (14%) | 99 (12%) |  |
| **Prior ICH** |  |  | 0.50 |
|  | 165 (9%) | 67 (8%) |  |

Most diagnoses were recorded as yes/no responses, unless otherwise indicated. Pre-stroke medical history of smoking and liver disease had missing responses for over 100 participants.

Of the 2,539 patients, 361 had follow-up data for 12-month mRS (Figure 1). Of these patients, 299 (83%) had a good functional outcome, and 62 (17%) had a poor functional outcome (Table 3). Patients with a pre-stroke diagnosis of HTN or dementia were more likely to have a poor functional outcome (p-value = 0.002 and p-value<0.001, respectively) (Table 4).

**Table 3:**
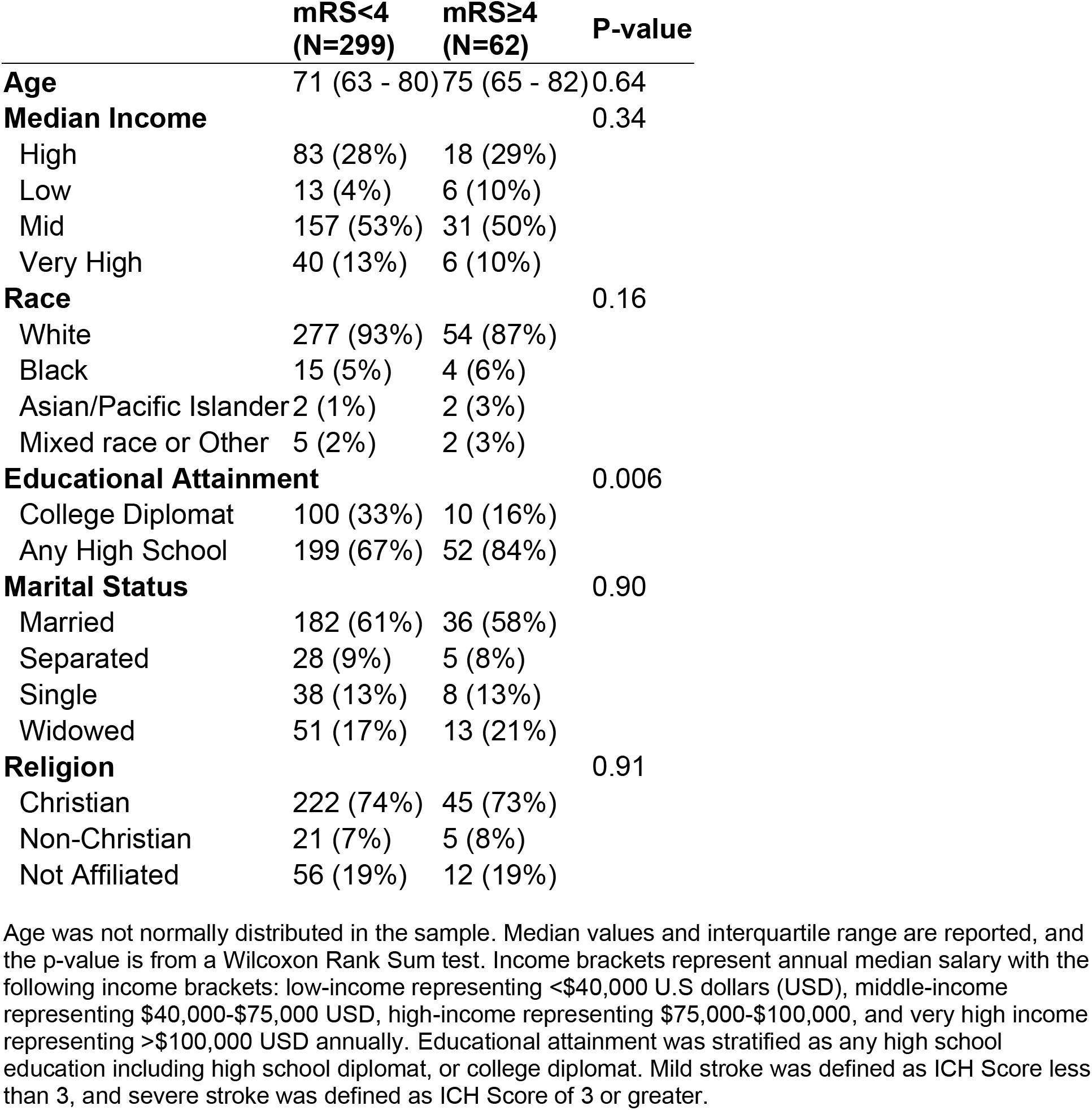
SDOH and 12-month functional outcome

|  | **mRS<4**<br>**(N=299)** | **mRS≥4**<br>**(N=62)** | **P-value** |
| --- | --- | --- | --- |
| **Age** | 71 (63 - 80) | 75 (65 - 82) | 0.64 |
| **Median Income** |  |  | 0.34 |
| High | 83 (28%) | 18 (29%) |  |
| Low | 13 (4%) | 6 (10%) |  |
| Mid | 157 (53%) | 31 (50%) |  |
| Very High | 40 (13%) | 6 (10%) |  |
| **Race** |  |  | 0.16 |
| White | 277 (93%) | 54 (87%) |  |
| Black | 15 (5%) | 4 (6%) |  |
| Asian/Pacific Islander | 2 (1%) | 2 (3%) |  |
| Mixed race or Other | 5 (2%) | 2 (3%) |  |
| **Educational Attainment** |  |  | 0.006 |
| College Diplomat | 100 (33%) | 10 (16%) |  |
| Any High School | 199 (67%) | 52 (84%) |  |
| **Marital Status** |  |  | 0.90 |
| Married | 182 (61%) | 36 (58%) |  |
| Separated | 28 (9%) | 5 (8%) |  |
| Single | 38 (13%) | 8 (13%) |  |
| Widowed | 51 (17%) | 13 (21%) |  |
| **Religion** |  |  | 0.91 |
| Christian | 222 (74%) | 45 (73%) |  |
| Non-Christian | 21 (7%) | 5 (8%) |  |
| Not Affiliated | 56 (19%) | 12 (19%) |  |

Age was not normally distributed in the sample. Median values and interquartile range are reported, and the p-value is from a Wilcoxon Rank Sum test. Income brackets represent annual median salary with the following income brackets: low-income representing <\$40,000 U.S dollars (USD), middle-income representing \$40,000-\$75,000 USD, high-income representing \$75,000-\$100,000, and very high income representing >\$100,000 USD annually. Educational attainment was stratified as any high school education including high school diplomat, or college diplomat. Mild stroke was defined as ICH Score less than 3, and severe stroke was defined as ICH Score of 3 or greater.

**Table 4:** Clinical characteristics and12-month functional outcome

|  | **mRS<4**<br>**(N=299)** | **mRS≥4**<br>**(N=62)** | **P-value** |
| --- | --- | --- | --- |
| **ICH Severity** |  |  | < 0.0001 |
| Mild | 229 (77%) | 36 (58%) |  |
| Severe | 22 (7%) | 17 (27%) |  |
| **Sex** |  |  | 0.21 |
| Female | 136 (45%) | 34 (55%) |  |
| **Smoking** |  |  | 0.071 |
| Current | 26 (9%) | 9 (15%) |  |
| History | 153 (51%) | 23 (37%) |  |
| Never | 112 (37%) | 29 (47%) |  |
| Missing | 8 (3%) | 1 (2%) |  |
| **Diabetes** |  |  | 0.60 |
|  | 61 (20%) | 14 (23%) |  |
| **Coronary Artery Disease** |  |  | 0.58 |
|  | 51 (17%) | 12 (19%) |  |
| **Atrial Fibrillation** |  |  | 0.58 |
|  | 57 (19%) | 9 (15%) |  |
| **Liver Disease** |  |  | 0.23 |
|  | 16 (5%) | 6 (10%) |  |
| **Dementia** |  |  | 0.0006 |
|  | 16 (5%) | 12 (19%) |  |
| **Prior Hemorrhage** |  |  | 0.29 |
|  | 34 (11%) | 10 (16%) |  |

Most diagnoses were recorded as yes/no responses, unless otherwise indicated. Pre-stroke medical history of smoking had missing responses for over 100 participants.

### Primary outcome analysis

Table 1 summarizes the SDOH by stroke severity. In univariate analysis, the only SDOH that were shown to have an association with stroke severity were annual median income and educational attainment were associated with stroke severity (p-value <0.0001 for both). Patients from low- and middle-income brackets comprised 66% of those with severe strokes while those with high- or very high-income brackets comprised 31%. In comparison, those from low- and middle-income brackets comprised 62% of mild strokes while those with high- or very high-income brackets comprised 34% of all mild strokes. College diplomates comprised only 8% of severe strokes and 18% of mild strokes, compared to those with any high school education who comprised 92% and 82% of severe and mild strokes respectively. Religion, race, and marital status were not significantly different between those with severe and mild stroke.

A multivariable logistic regression model adjusted for age, education level, income level, and pre-stroke diagnosis of CAD revealed a significant adjusted association for education level alone (OR 2.37 95CI 1.77, 3.19).

### Secondary outcome analysis

Table 3 summarizes SDOH and stroke severity by 12-month mRS. Among the 361 patients with 12-month mRS, 62 individuals (17%) had poor functional outcomes. Those with severe strokes and lower education level were more likely to have poor outcomes than those with mild strokes or those with higher education level. Median income, religion, race, and marital status were not significantly different between those with good versus poor functional outcome (Table 3). Additionally, pre-stroke diagnoses of either HTN and dementia were also associated with poor functional outcomes (Table 4).

After adjustment for stroke severity, education level, and a pre-stroke diagnosis of HTN and dementia, high school-only education was associated with a 57% reduction in likelihood of a good functional outcome at 12-months post-ICH, compared to college education (OR 0.43 95CI 0.19, 0.93). Patients with a pre-stroke diagnosis of HTN were 75% less likely to recover to a good outcome than those who did not (OR 0.25, 95CI 0.09, 0.67) when controlling for stroke severity, dementia, and education. Those with a pre-stroke diagnosis of dementia are 80% less likely to recover to a good outcome those who did not have a diagnosis of dementia (OR 0.20, 95CI 0.08, 0.54) when controlling for stroke severity, education, and HTN.

## Discussion

Our results indicate that pre-stroke educational attainment is an independent predictor of hemorrhagic stroke severity and subsequent functional recovery at 12-months post-stroke, emphasizing that specific social determinants of health can impact severity of stroke and stroke recovery. Despite decades of investigative trials and research in hemorrhagic stroke, ours if the first to give attention to educational attainment in this stroke population. The novel findings that education impacts both hemorrhagic stroke severity and functional outcome implies two potential mechanisms. First, educational attainment could be a surrogate marker for other social determinants that impact overall health; for example, those with lower education likely had less exposure or access to preventative health services or had had more exposures to environments that worsened traditional risk factors. Similarly, those with higher education may have had more exposure to health services through their life course or had more exposures to environments that decreased traditional risk factors. Previous studies have generated theoretical models for how SDOH, such as education, can serve independent causal factors for stroke^9,18^.

A second proposed mechanism is that education is both a social and biological determinant. In the theory proposed by Umarova et al., education could act a surrogate maker for cognitive reserve that mediates the relationship between injury and recovery^6^. These authors also suggest that in those with higher education, the higher conferred cognitive reserve could not only be protective but advantageous in recovery, especially when rehabilitation focuses on learning behavioral strategies to recruit new networks^6^. Further supporting this theory, is our finding that those with a pre-stroke diagnosis of dementia, and presumably less cognitive reserve, were 80% less likely to recover to a good functional outcome. This theoretical mechanism of cognitive reserve could partially explain disparate outcomes in hemorrhagic stroke survivors described in prior studies which failed to examine educational attainment but did not find other social determinants, such as race, to be associated with outcomes^11^.

Similarly congruent with our findings, a recent large, international prospective cohort study across 21 diverse countries also found that in 155,722 patients, low education was more predictive of all-cause mortality than hypertension, and contributed just as much, if not more, attributable risk for both cardiovascular and stroke events than tobacco use^3^. Another recent analysis of ischemic stroke patients found larger strokes did not lead to as severe cognitive or functional decline in those patients with higher levels of educations compared to those with lower levels of education, even when accounting for age^6^.

Our first finding that educational attainment is associated with stroke severity illustrates the Healthy People 2030 position that factors other than traditional health indicators are just as impactful to health outcomes. Given the strong link of social determinants and outcomes across health conditions, we hope that our second finding that educational attainment is also associated with functional outcome is the motivation to restructure stroke prevention education and stroke recovery education. Clinical trials of education and information tailored to the educational attainment of the patient will be needed to determine whether such interventions could ultimately lead to a reduction of disparities in stroke morbidity and mortality.

### Limitations

The limitations of this study are the relatively homogenous population of patients admitted to MGH. Our cohort represents an older and more racially homogenous population than previously studied stroke cohorts^9,11,12,19,20^. However, even in this population, educational attainment played a role in the severity and recovery from ICH. Next, a notable finding in our results was that a pre-stroke diagnosis of HTN was not an independent predictor of stroke severity. The limitation of our measure is that HTN is a dichotomous yes/no measure and does not capture the severity or duration of a patient’s hypertension. Lastly, we have significant attrition of our sample for the 12-month mRS that likely does not represent random loss to follow up. Withdrawal of care was also not investigated in this study but remains a challenge in studying long term recovery due to survivor bias.

Future studies should use more diverse populations to investigate the impact of educational attainment on stroke severity. Next, while we examined level of education, future studies should examine if education quality has similar impacts on stroke, as stated by the Healthy People 2030 framework. Additionally, more detailed assessments of wellbeing could also provide key information to factors that promote recovery; while we used the mRS, this crude clinical score may not capture the extent of impact that social determinants, such as education, have on stroke recovery. Lastly, given that the link between SDOH and stroke is well-established, future studies should focus on systems-level interventions for patients with low education to help with recovery.

## Conclusion

Our findings show the importance of educational attainment in ICH severity and recovery. We found that education was an independent predictor of ICH severity, and lower levels of education were associated with worse functional outcomes 12 months after initial stroke. Our findings suggest that a generalized approach to stroke education may be insufficient and ineffective. Educational attainment may act as both a social and a biological determinant, but regardless, could represent a modifiable risk factor for ICH. With these results, we hope future studies will investigate the impact of providing patient-centered information that accounts for pre-stroke educational levels for hemorrhagic stroke prevention and recovery.

## Data Availability

The participants of this study did not give written consent for their data to be shared publicly, so due to the sensitive nature of the research supporting data is not available.

## Acknowledgments

Drs. Yechoor and Rosand had full access to all the data in the study and take responsibility for the integrity of the data and the accuracy of the data analysis.

*Study concept and design: Yechoor, Rist, Rosand*

*Acquisition, analysis, and interpretation of data: Yechoor, Rist, Rosand*

*Drafting of the manuscript and critical revisions for important intellectual content:*

*Yechoor, Rist, Ganbold, Kourkoulis, Mora, Mayerhofer, Parodi, Anderson, Rosand*

*Statistical analysis: Yechoor, Rist, Mora*

